# Prediction of COVID-19 Pandemic of Top Ten Countries in the World Establishing a Hybrid AARNN LTM Model

**DOI:** 10.1101/2020.12.31.20249105

**Authors:** Padmabati Gahan, Monalisha Pattnaik, Agnibrata Nayak, Monee Kieran Roul

## Abstract

The novel COVID-19 global pandemic has become a public health emergency of international concern affecting 215 countries and territories around the globe. As of 28 November 2020, it has caused a pandemic outbreak with a total of more than 6,171,5119 confirmed infections and more than 1,44,4235 confirmed deaths reported worldwide. The main focus of this paper is to generate LTM real-time out of sample forecasts of the future COVID-19 confirmed and death cases respectively for the top ten profoundly affected countries including for the world. To solve this problem we introduced a novel hybrid approach AARNN model based on ARIMA and ARNN forecasting model that can generate LTM (fifty days ahead) out of sample forecasts of the number of daily confirmed and death COVID-19 cases for the ten countries namely USA, India, Brazil, Russia, France, Spain, UK, Italy, Argentina, Colombia and also for the world respectively. The predictions of the future outbreak for different countries will be useful for the effective allocation of health care resources and will act as early-warning system for health warriors, corporate leaders, economists, government/public-policy makers, and scientific experts.

## 1. Introduction

The COVID-19 global pandemic is overwhelming hospitals and emergency response systems in many countries like, USA, India, Brazil, Russia, France, Spain, UK, Italy, Argentina, Colombia in and is expected to influence provision of care years to come. Health + Hospitals (H+H) have played a crucial role in entire globe’s response to COVID-19. As more than a billion public health care system with more than billion acute-care hospitals and post-acute care facilities, global Health + Hospitals (H+H) has played a significant role in responding to the COVID-19 pandemic outbreak, particularly for the countries like USA, India, Brazil, Russia, France, Spain, UK, Italy, Argentina, Colombia and also for the World. We describe our experience developing a forecasting model of COVID-19 confirmed and death cases for top ten profoundly affected countries including world dataset.

In December 2019, Wuhan city of China became the hub of an outbreak of pneumonia of mysterious cause, latter named as COVID-19, which raised intense attention not only within China but globally (Chakraborty et al., 2020, Guan et al., 2020, Ordonez et al., 2019 and Jung et al., 2020). The COVID-19 pandemic is the most significant global crisis that affected almost all the countries of the globe (Boccaletti et al., 2020). As of 28 Nov. 2020, an outbreak of COVID-19 has resulted in a total of more than 6,171,5119 confirmed infections and more than 1,44,4235 confirmed deaths reported worldwide (Nishiura et al., 2020). On March 11, WHO publicly characterized COVID-19 as a “global pandemic”, and shortly after that, the United States declared COVID-19 outbreaks a national emergency. The COVID-19 has caused a great threat to the health and safety of people all over the world due to its widespread and prospective harm. Thus, the studies of the novel COVID-19 pandemic and its future development trend has become a state-of-the-art research topic at this moment. We are therefore stimulated to ask: Can we generate real-time forecasts of daily new COVID-19 confirmed and death cases for the top ten profoundly affected countries namely USA, India, Brazil, Russia, France, Spain, UK, Italy, Argentina, Colombia and also for the World? To answer this question, we study classical and modern forecasting techniques for which the prediction accuracy largely depends on the availability of data (Petropoulos et al., 2020). In outbreaks of COVID-19 pandemic, there are limited data available, making predictions generally uncertain. From previous studies, it was evident that the timing and location of the outbreak facilitated the rapid transmission of the virus within a highly mobile population (Roosa et al., 2020, Boldog et al., 2020 and Russel et al., 2020). In most of the affected countries, the governments implemented a strict lockdown in subsequent days of initial transmission of the virus and within hospitals; patients who fulfil clinical and epidemiological characteristics of COVID-19 are immediately quarantined. The constant increase in the global number of COVID-19 confirmed cases is putting a substantial burden on the health care system for these top ten countries. To anticipate additional resources to combat the pandemic, various mathematical and statistical forecasting tools (Li et al., 2020 and Wu et al., 2020) and outside China (Fanelli et al., 2020, Kucharski et al., 2020) were applied to generate short-term and long-term forecasts of reported cases. These model predictions have shown a wide range of fluctuations. Since the time series datasets of COVID-19 confirmed and death cases contain nonlinearity, nonstationary and non-Gaussian patterns, therefore, making decisions based on an individual model would be critical. In this study, we propose a hybrid modeling approach to generate LTM out-of-sample forecasts for multiple countries. In traditional time series forecasting, the ARIMA model is used predominantly for forecasting linear time series (Box et al., 2015). Machine learning models such as ANN, ARNN and WBF (Paul a al., 2017, Nury et al., 2017, Aminghafari et al., 2007, Beraouda et al., 2006 and Fay et al., 2007) are proved to perform well for nonlinear data structures in time series data. COVID-19 data sets are neither purely linear nor nonlinear. They usually contain both linear and nonlinear patterns. If this is the case, then the individual ARIMA or ARNN or ANN or WBF is not enough to model such situations. Therefore, the combination of linear and nonlinear models can be well suited for accurately modeling such complex autocorrelation structures (Percival et al., 2020 and Chakraborty et al, 2020) which reduce the bias and variances of the prediction residuals of the component models. Several hybrid methodologies were discussed in literature to solve a variety of time series problems arose in economic parameters, economic growth, GDP and wind speed (Tumer et al., 2018, Kordanuli et al. 2017, Khashei et al., 2011, Milacic et al., 2017, Cadenas et al., 2010, Maleki et al. 2018 and Chakraborty et al., 2019, James et al., 2013 and Pattnaik et al. 2018). Zhang’s hybrid ARIMA-ANN model (Zhang et al., 2003) has gained popularity due to its capacity to forecast complex time series accurately. The pitfall of hybrid ARIMA-ANN model lies in the selection of the number of hidden layers in the ANN architecture that involves subjective judgment. To ignore this drawback, we took recourse to the ARNN model approach. ARNN fits a feed-forward neural network model with only one hidden layer to a time series with lagged values of the series as inputs. The advantage of ARNN over ANN and WBF is that ARNN is a nonlinear autoregressive model, and it provides less complexity, easy interpretability and better prediction as compared to others in many situations. Due to this, ARNN is getting more attention in recent literature of non-stationary time series forecasting (Hydmann, 2018). In the absence of vaccines or antiviral drugs for COVID-19, these estimates will present an insight into the resource allocations for the extremely influential countries to keep this global pandemic under control. Besides detaching light on the dynamics of COVID-19 dissemination, the practical intent of this real-time data-driven analysis is to provide health warriors, corporate leaders, economists, government/public-policy makers, and scientific experts with realistic estimates for the magnitude of the pandemic for policy-making. The main inspiration of this paper is to handle the dynamism of the characteristics of the time series COVID-19 data. Taking a final decision in policy making based on a component model may be risky in such a severe problem like COVID-19 pandemic confirmed and death cases forecasting where one frequently observes changes in the dynamic properties of the variable being measured. Hybridization of two or more models is the most common solution to this problem (Oliveira et al., 2014). Even from the practitioners’ point of view, hybrid models are more effective when the complete data characteristics are not known (Kuncheva, 2004). Motivated from these discussions, this paper proposes a novel hybrid ARIMA-ARNN (AARNN) model that captures complex data structures and linear plus nonlinear behavior of COVID-19 data sets. In the first phase of our proposed model, ARIMA catches the linear patterns of the data set. Then the ARNN model is employed to capture the nonlinear patterns in the data using residual values obtained from the base ARIMA model. The proposed model has easy interpretability, robust predictability and can adapt seasonality indices as well. Through experimental evaluation, we have shown the excellent performance of the proposed hybrid AARNN model for the COVID-19 global pandemics forecasting for profoundly affected top ten countries including world data sets.

The rest of the paper is structured as follows: In Section 2, country-wise COVID-19 confirmed and death cases datasets, preliminary of data analysis and performance evaluation metrics are presented. In Section 3, we discuss the proper formulation of the proposed hybrid AARNN model. We discuss the experimental evaluations and results for LTM out-of-sample forecasts of COVID-19 global pandemic for the top ten countries including for the world in Section 4. The discussions about the results and practical implications are presented in Section 5. Finally, in Section 6, concluding remarks of this study are presented.

## 2. Data and preliminary analysis

We explore on the daily confirmed and death cases of COVID-19 pandemic for ten countries namely USA, India, Brazil, Russia, France, Spain, UK, Italy, Argentina, Colombia and for the World respectively. The datasets are retrieved by the Global Change Data Lab^1^. The starting date of all the countries is different but the ending date is same for all i.e. November 28, 2020. Present section includes the brief discussion about the datasets, the development of the proposed hybrid model and finally, the application of the proposed model to generate long-term-memory (LTM) out-of-sample forecasts of the COVID-19 confirmed and death cases for the above ten countries including the world.

### 2.1 COVID-19 datasets

Eleven univariate time series datasets of both confirmed and death cases of COVID-19 are collected for the real-time prediction purpose for the ten countries including the world. Different earlier studies have forecasted future COVID-19 confirmed cases only for different countries using mathematical and traditional time series models [].We try to investigate accurate prediction both of COVID-19 confirmed and death cases of top ten profoundly affected countries. Initial day of daily laboratory-confirmed and –death cases for all the countries are different but the last day of observation for all the countries is considered same timeline i.e. 28.11.2020. Table 2 shows the description of COVID-19 data sets of confirmed cases and death cases for USA, India, Brazil, Russia, France, Spain, UK, Italy, Argentina, Colombia and for world respectively. The dataset of confirmed cases for the USA, India, Russia, France, Spain, UK and world contains a total of more than 300 observations whereas the dataset for Brazil, Argentina and Colombia contains less than 300 observations. The dataset of death cases for all the studied countries are in between a total of 205 and 300 observations except for world.

**Table 1:**
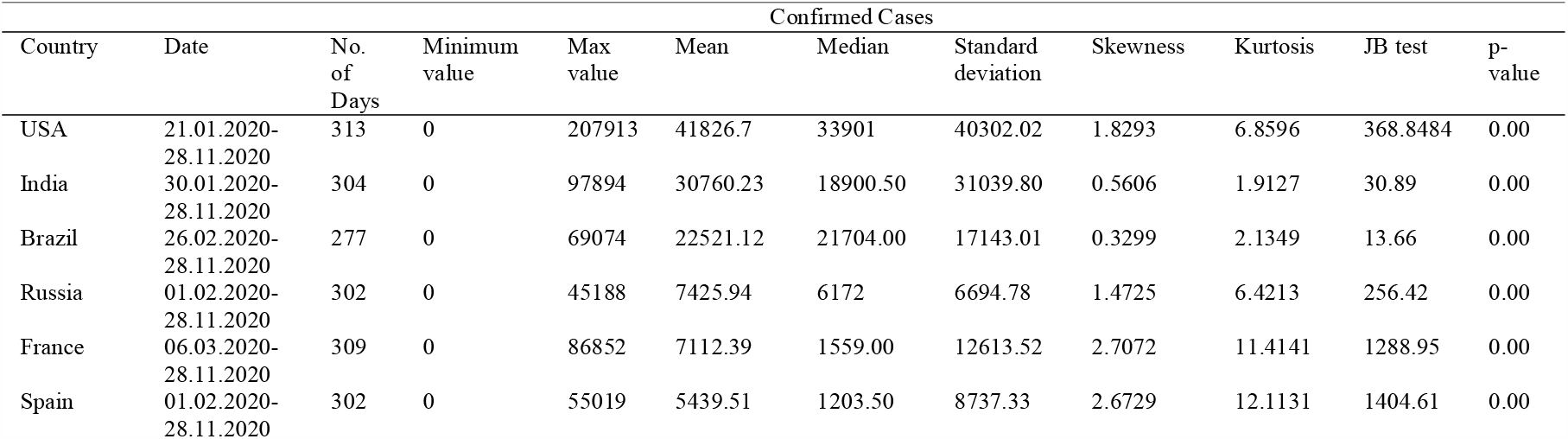
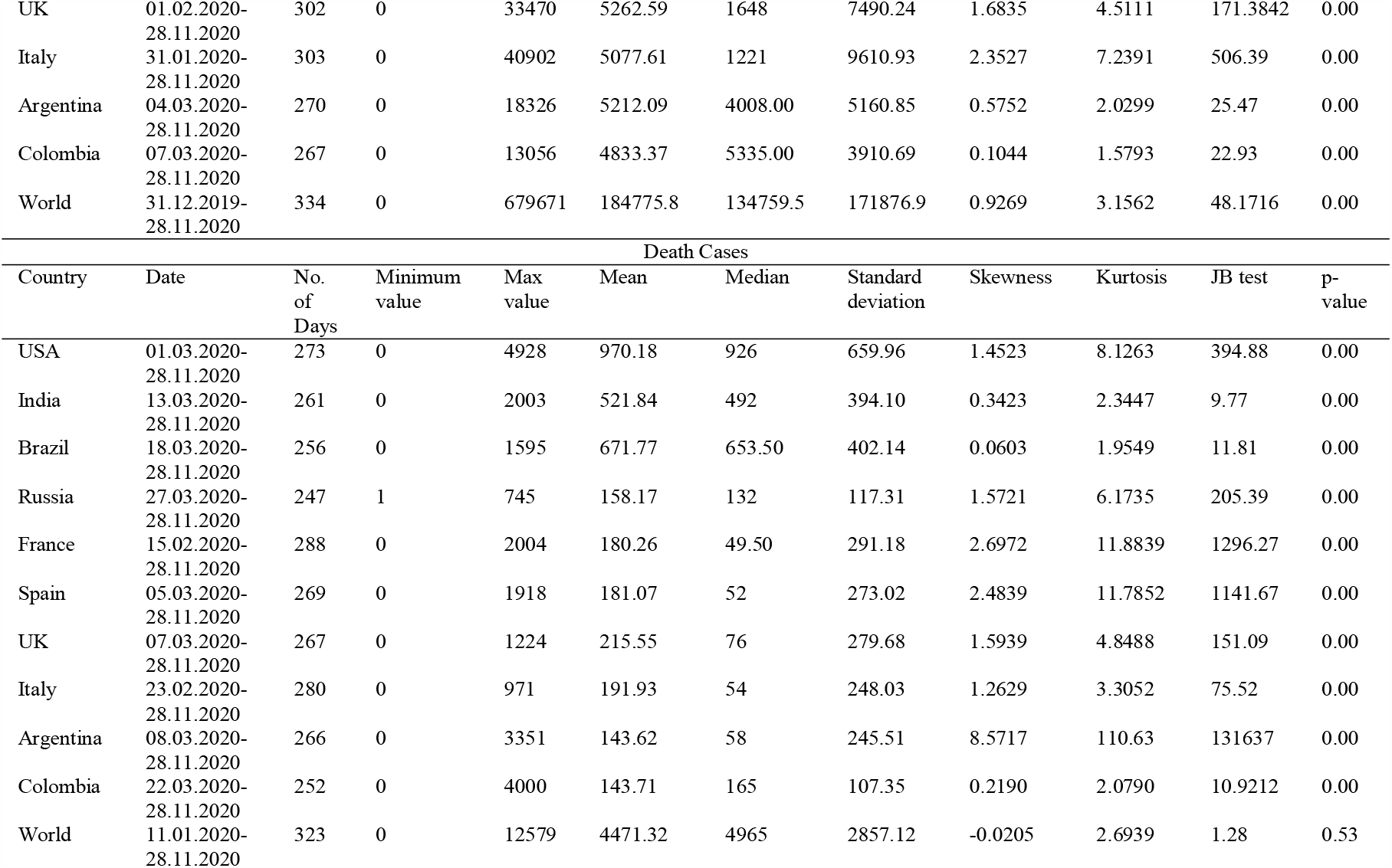
Descriptions of COVID-19 data sets of confirmed cases and death cases for USA, India, Brazil, Russia, France, Spain, UK, Italy, Argentina, Colombia and World respectively

**Table 2:** Algorithm of Proposed Novel Hybrid AARNN Model

| Algorithm of proposed novel hybrid AARNN model |  |
| --- | --- |
| 1. | Given a time series of length $n$, input the in-sample (training) COVID-19 daily confirmed cases and death data. |
| 2. | Determine the theta ($p, d, q$) model using the in-sample (training) data.<br>• ARIMA parameters $p, d, and q$ values are selected using the procedures described in Section 3.1.<br>• Obtain the predictions using the selected ARIMA ($p, d, q$) model for the in-sample data and generate required number of out-of-sample forecasts.<br>• Obtain the residual series ($\varepsilon_t$) by subtracting ARIMA predicted values from the original training series. |
| 3. | Determine the best ARNN ($p, k$) model on the training residual series($\varepsilon_t$) generated by ARIMA model.<br>• Perform lag selection for the training of residual series and apply ARNN model with $p$ selected lagged inputs from the residual series($\varepsilon_t$) and $k$ hidden units in one layer as described in Section 3.2.<br>• Obtain predictions using the ARNN model ($\hat{\varepsilon}_t$). |
| 4. | Final predictions ($\hat{Z}_t$) are obtained by combining the ARIMA predictions with ARNN predictions ($\hat{\varepsilon}_t$). |

### 2.2 Preliminary data analysis

A summary of the COVID-19 data sets of confirmed and death cases are shown in Table 1. From p-values it indicates that all the eleven COVID-19 data sets of both confirmed and death cases are not normally distributed. Table 3 and 4 (A-1 & A-2) show the daily confirmed and death cases of COVID-19 pandemic of ten countries including world respectively. The epidemic curves still not showing the spiky declining nature. We bound our attention for trend and non-seasonal models. We consider a novel approach which assumes that the trend will tend to infinity in the future. To fit the traditional ARIMA model, hybrid ARIMA-ANN, WARIMA and proposed AARNN models of COVID-19 confirmed and death cases, the parameters namely *p* and *q* are specified by plotting ACF and PACF plots (see Table 3 and 4).

**Table 3:**
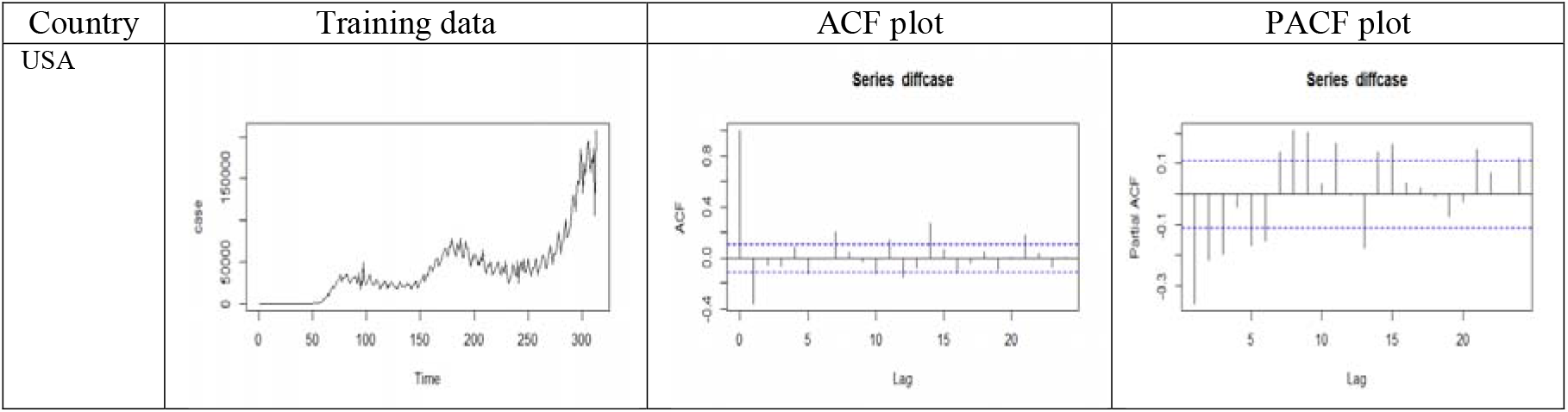
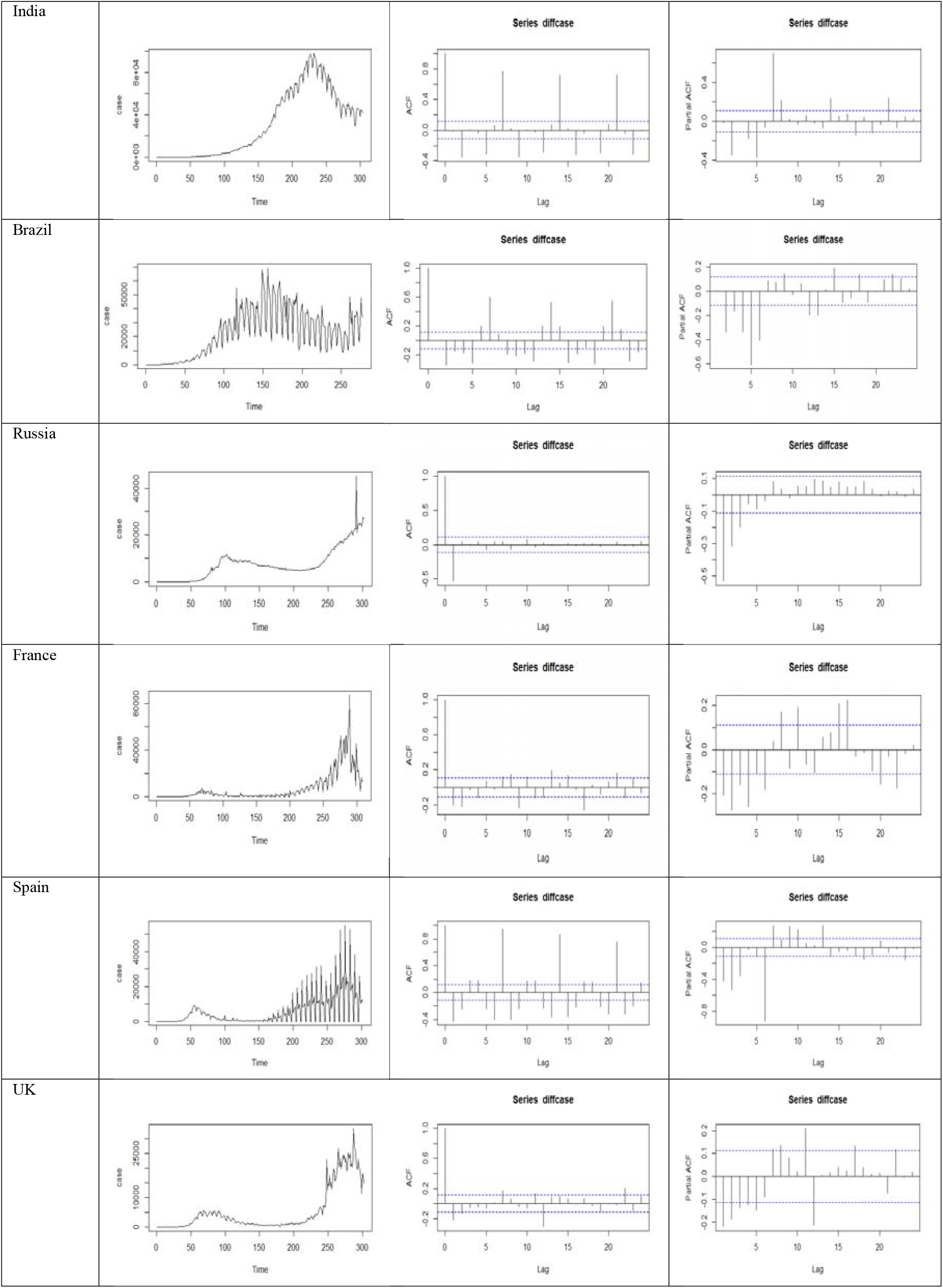
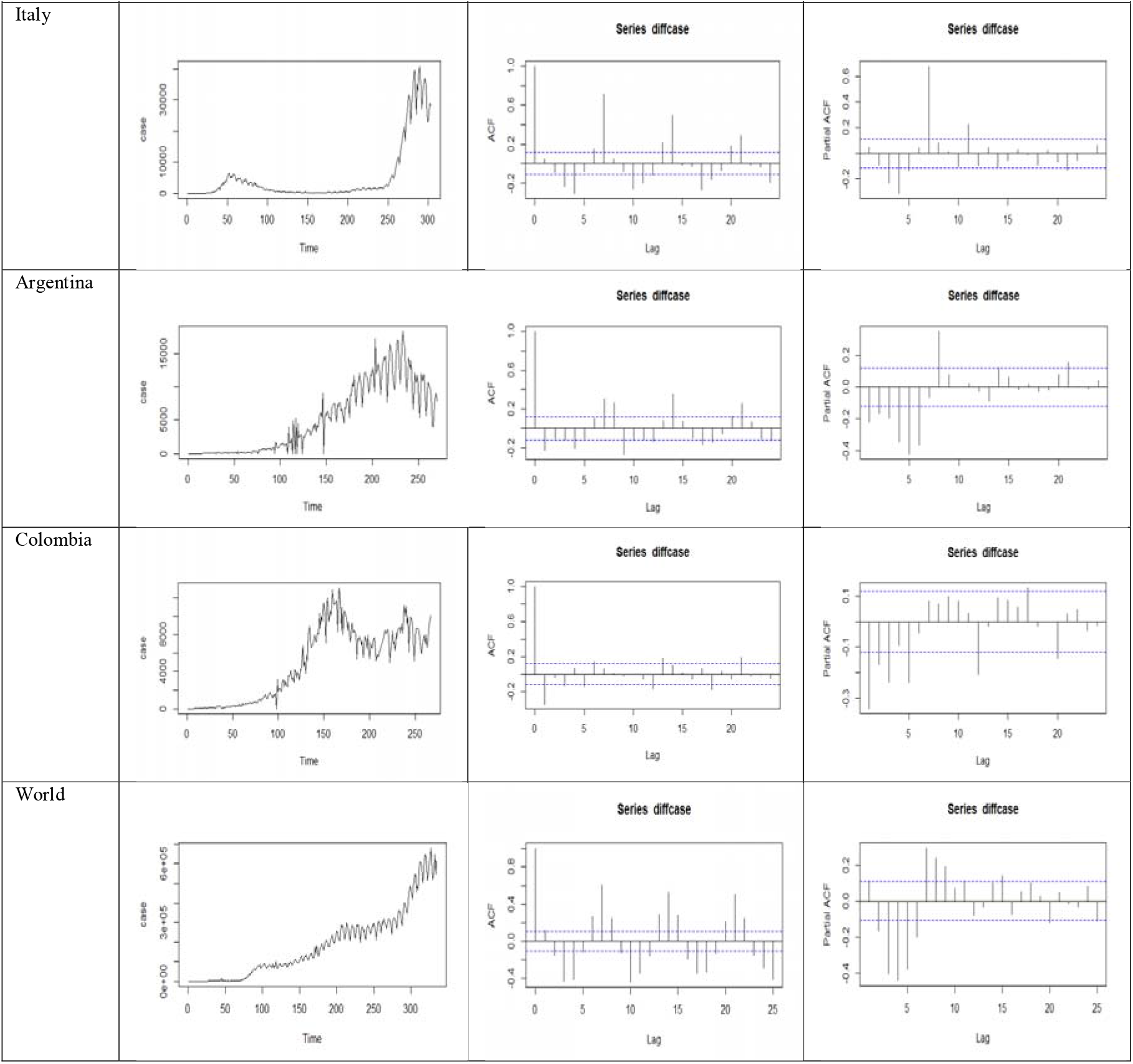
Training COVID-19 data sets of confirmed cases and corresponding ACF, PACF plots for USA, India, Brazil, Russia, France, Spain, UK, Italy, Argentina, Colombia and World

**Table 4:**
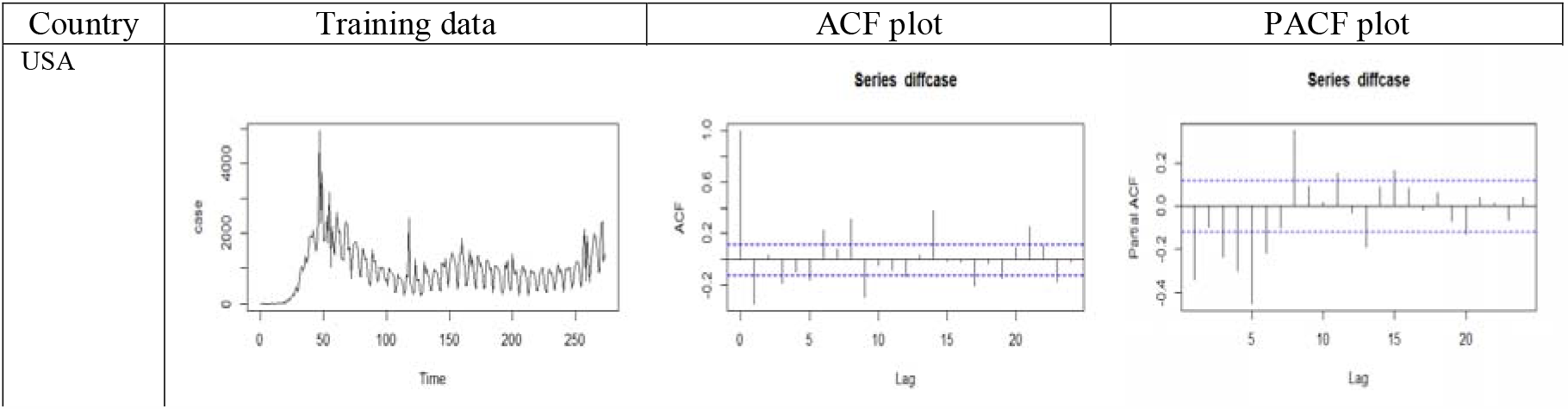
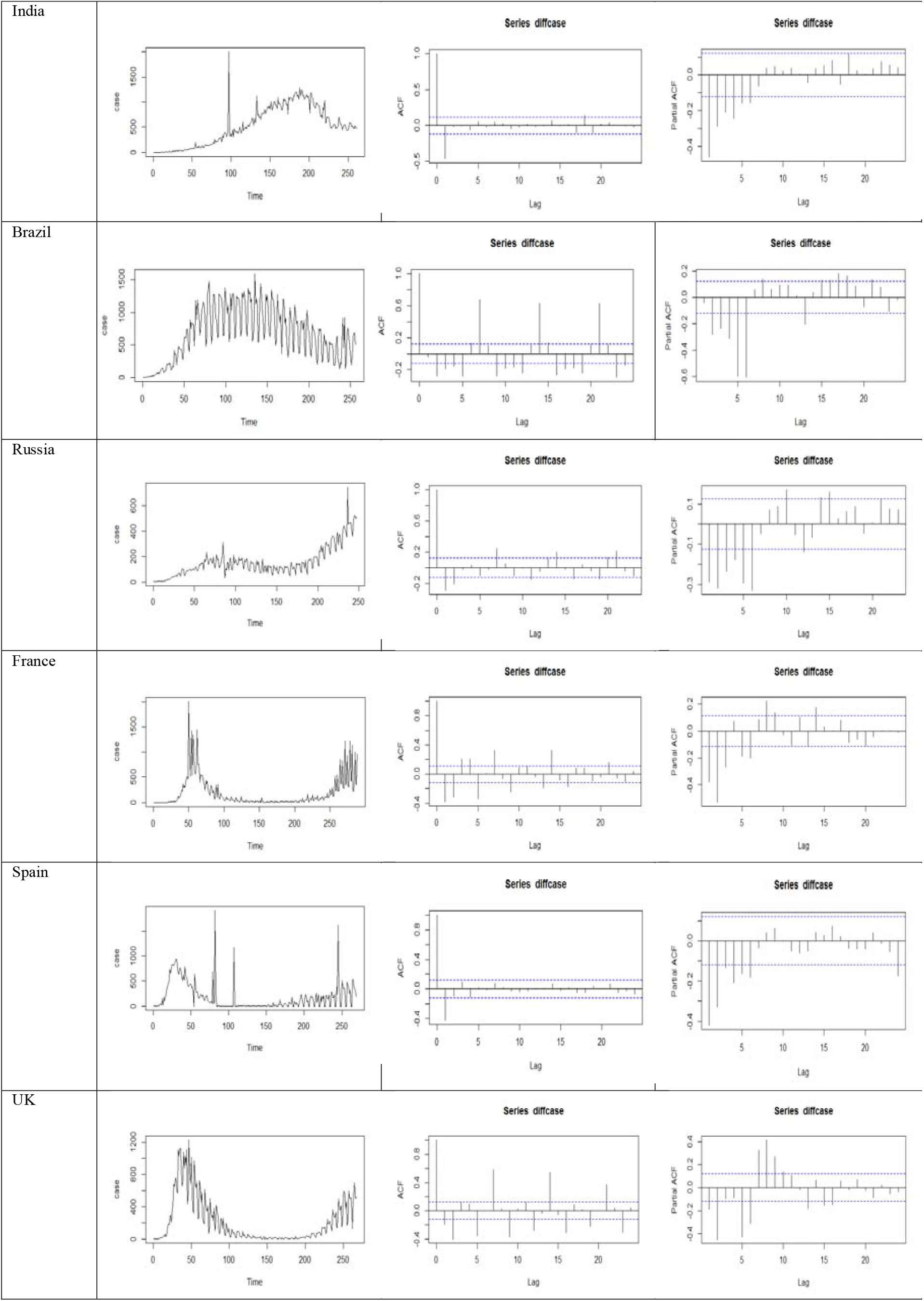
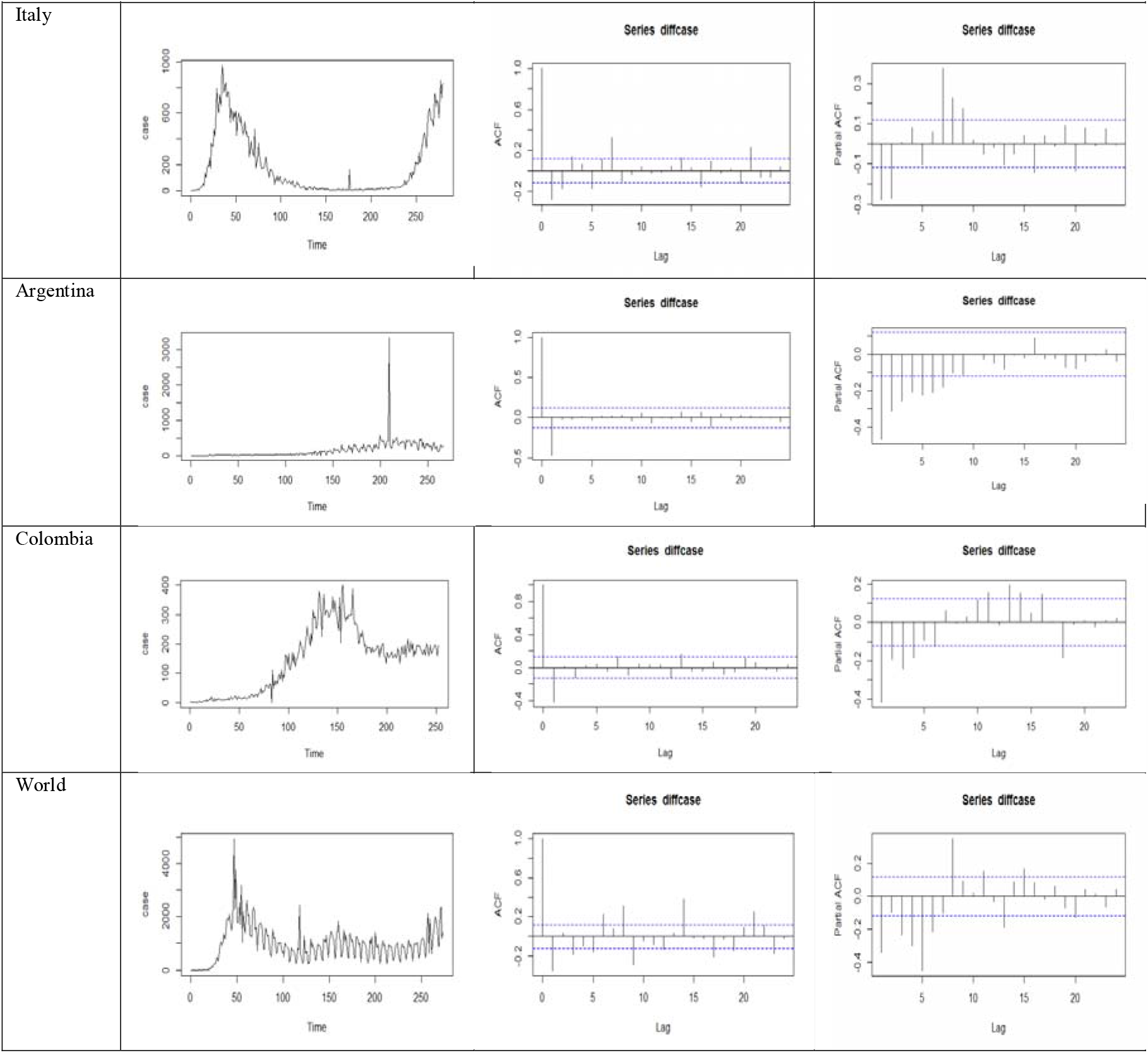
Training COVID-19 data sets of death cases and corresponding ACF, PACF plots for USA, India, Brazil, Russia, France, Spain, UK, Italy, Argentina, Colombia and World

### 2.3 Performance evaluation metrics

The performance of different forecasting models is evaluated based on RMSE and MAE for the COVID-19 data sets (Ahmed et al., 2010 and Chakraborty et al., 2020). The mathematical expressions of these performance evaluation metrics are as follows.

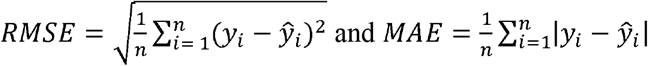

where, *y*_*i*_ is the actual output, *ŷ*_*i*_ is the predicted output, and denotes the number of data points. By definition, the lower the value of these performance metrics, the better is the performance of the concerned forecasting model.

## 3. Methodology

To forecast COVID-19 confirmed and death cases, we adopt hybrid time series forecasting approaches combining ARIMA and ARNN techniques. The proposed AARNN hybrid model overcomes the deficiencies of the individual linear and nonlinear and other hybrid time series models. Before describing the methodology, we give a brief description of the individual traditional and advanced models to be used in the hybridization.

### 3.1 ARIMA Model

The autoregressive integrated moving average (ARIMA) model, developed by (Box and Jenkin 2015), is a linear regression model indulged to eliminate linear tendencies in stationary time series data. The model is expressed as ARIMA (p,d,q) where p, d, and q are integer parameter values that decide the structure of the model. More specifically, p and q are the order of the AR model and the MA model respectively, and parameter d is the level of differencing applied to the time series data. The mathematical expression of the ARIMA model (Chakraborty et al. 2019) is as follows:

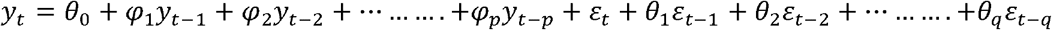

where *y*_*t*_ is the actual value, *ε*_*t*_ is the random error at time t, *φ*_*i*_ and *θ*_*j*_are the coefficients of the model.

It is assumed that *ε*_*t−1*_ (*ε*_*t−1*_ = *y*_*t−1* −_ *ŷ* _*t−1*_)has zero mean with constant variance, and satisfies the i.i.d condition. The methodology consists of four iterative steps:

1. Model identification and model selection
2. Parameter estimation of the model parameters
3. Model diagnostics checking (namely, residual analysis) are performed to find the ‘best’ fitted model
4. Model forecasting

In the model identification and model selection step, differencing is applied once or twice to achieve stationarity for non-stationary data. As stationarity condition is satisfied, the ACF plot and the PACF plot are examined to select the AR and MA model types. The parameter estimation step involves an optimization process utilizing metrics such as the AIC and/or the BIC. Finally, in the model checking step, the residual analysis is carried out to finalize the ‘best’ fitted ARIMA model. ARIMA model is a data-dependent approach that can adapt to the structure of the data set. But it has the major disadvantage that any significant nonlinear data and non-Gaussian set can restrict the ARIMA model. Therefore, some individual and hybrid models uses ARNN, ANN and WBF models to deal with the nonlinear data patterns for forecasting intricate time series structure.

### 3.2 ARNN Model

ANN is a widely used supervised learning model, highly useful for sophisticated nonlinear time series forecasting. Neural nets are based on simple mathematical models of the brain, used for intricate nonlinear forecasting. Any neural net architecture can be described as a network of “neurons”, arranged in layers, namely the input layer, hidden layer, and output layer. The information from one layer to another layer is passed using weights that are selected using a risk minimization based ‘learning algorithm’ (Chakraborty et al. 2020). The ARNN model is a modified neural network model especially designed for time series datasets which uses a pre-specified number of hidden neurons in its architecture (Faraway and Chatfield 1998). It uses lagged values of the time series as inputs to the model. ARNN (*p, k*) is a nonlinear feed-forward neural net model with one hidden layer (having *p* lagged inputs) and *k* hidden units 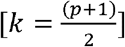 for non-seasonal data in one hidden layer (Hyndman and Athanasopoulos 2018). It also uses AIC as the criterion for comparing different models created by ARNN. Here *ŷ*_*t*_ is computed using selected past observations *y*_*t*−1_, *y*_*t*−2_, …… *y*_*t−p*_ as the inputs. Weights of the ARNN model are trained using a gradient descent back propagation algorithm (Rumelhart et al. 1985).

### 3.3 Proposed AARNN Model

ARIMA model is one of the traditional statistical models for linear time series prediction. On the other hand, the advanced ARNN model can capture nonlinear trends in the data set. So, the two models are consecutively combined to encompass both linear and nonlinear tendencies in the model (Aladag et al., 2009). A hybrid strategy that has both linear and nonlinear modeling abilities is a good alternative for forecasting dengue cases. Both the ARIMA and the ARNN models have different capabilities to capture data characteristics in linear or nonlinear domains. Thus, the hybrid approach can model linear and nonlinear patterns with improved overall forecasting performance. There exists numerous time series models in the literature, and several research shows forecast accuracy improves in hybrid models. The aim of developing a novel hybridization is to harness the advantages of individual models and reduce the risk of failures of these models. The underlying assumption of the hybrid approach based on linear and nonlinear model assumption is that the relationship between linear and nonlinear components are additive. The strength of individual models for hybridization is very important, and this selection is essential to show the consistent improvement over single models. This paper presents a novel hybridization of ARIMA and ARNN (AARNN) to overcome the limitation of individual models and utilize their strengths. With the combination of linear and nonlinear models, the proposed methodology can guarantee better performance as compared to the component models.

The hybrid model (*Z*_*t*_) can be represented as follows:

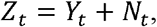

where *Y*_*t*_ is the linear part and *N*_*t*_ is the nonlinear part of the hybrid model. Both *Y*_*t*_ and *N*_*t*_ are estimated from the data set. Let, *Ŷ*_*t*_ be the forecast value of the ARIMA model at time t and *ε*_*t*_ represent the residual at time t as obtained from the ARIMA model; then. *ε*_*t*_ *= Z*_*t*_*−Ŷ*_*t*_

The residuals are modeled by the ARNN model and can be represented as follows

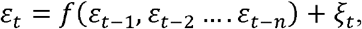

where f is a nonlinear function modeled by the ARNN approach and *ξ*_*t*_ is the random error. Therefore, the combined forecast is 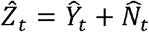,
where, 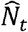 is the forecast value of the ARNN model. The rationale behind the use of residuals in the diagnosis of the sufficiency of the proposed hybrid model is that there is still autocorrelation left in the residuals which ARIMA could not model. This work is performed by the ARNN model which can capture the nonlinear autocorrelation relationship. In summary, the proposed hybrid AARNN model works in two phases. In the first phase, an ARIMA model is applied to analyze the linear part of the model. In the next stage, an ARNN model is employed to model the residuals of the ARIMA model. The hybrid model also reduces the model uncertainty which occurs in inferential statistics and forecasting time series. The algorithmic representation of the proposed hybrid AARNN model is also given in Table 2 (Chakraborty et al., 2020).

## 4. Experimental evaluations and results

It is a challenging task to study the characteristics and nature of the limited COVID-19 data sets. With the availability of COVID-19 datasets of both confirmed and death cases for ten different countries namely USA, India, Brazil, Russia, France, Spain, UK, Italy, Argentina and Colombia including world, we consider one significant problem relevant to ongoing COVID-19 global pandemic. This problem deals with the real-time data-driven forecasts of the daily COVID-19 confirmed and death cases of the above top ten profoundly affected countries. We propose a novel hybrid ARIMA-ARNN (AARNN) model to forecast daily COVID-19 confirmed and death cases of these countries. This proposed novel hybrid model can be treated as the best to notify the health practitioners, corporate leaders, economists, government/public-policy makers, and scientific experts to fight against the COVID-19 global pandemic.

Eleven univariate time series COVID-19 datasets of daily confirmed and death cases for USA, India, Brazil, Russia, France, Spain, UK, Italy, Argentina, and Colombia including world are considered for training the proposed novel hybrid AARNN model and the traditional (ARIMA), advanced (ARNN, ANN and WBF) and other hybrid models (ARIMA-ANN and WARIMA). The datasets are nonlinear, nonstationary, and non-Gaussian in nature and statistical tests confirmed this (see Section 2.2 and refer to Table 1). We experimentally evaluate the performances of ARIMA, ARNN, ANN, WBF, hybrid ARIMA-ANN, WARIMA model (Chakraborty et al., 2020), in comparison with our proposed novel hybrid AARNN model (Zhang, 2003 and Chakraborty et al., 2020) for all these eleven COVID-19 datasets. We have used RMSE and MAE, to evaluate the predictive performance of the models used in this study (James et al., 2013 and Chakraborty et al., 2020). Since the number of data points of COVID-19 pandemic datasets is limited thus implementing the advanced deep learning techniques will simply over-fit the datasets (Hastie et al., 2009 and Chakraborty et al., 2020).

We start the experimental evaluation for all the eleven datasets with the ARIMA model using ‘forecast’ (Hyndman et al., 2020) statistical package in R statistical software. We fit the ARIMA model using auto.arima function. As the ARIMA model is fitted, predictions are generated for 50-time steps ahead for LTM forecasts for all the eleven COVID-19 confirmed and death cases datasets respectively. We also compute training data predicted values and calculate the residual errors using the traditional individual model ARIMA model. Plots of the predicted and residual series of both confirmed and death cases of COVID-19 pandemic are given in Table 5 (A-3). As the ARIMA model is fitted on the residual time series, predictions of both confirmed and death cases are generated for the next fifty (29^th^ November 2020 to 17^th^ January 2021) time steps respectively. Further, both the ARIMA, ANN, WBF and proposed ARNN residual forecasts are added together to get the final out-of-sample forecasts of both confirmed and death cases COVID-19 outbreaks using the traditional hybrid ARIMA-ANN, WARIMA and the proposed hybrid AARNN model for the next fifty (29^th^ November 2020 to 17^th^ January 2021) days respectively. The proposed hybrid AARNN model for actual and fittings (training data) of COVID-19 confirmed and death cases for these ten countries including world data respectively are displayed in Table 6 (A-4). The real-time LTM out of sample forecasts (50 days ahead) of COVID-19 confirmed and death cases using ARIMA, ARNN and proposed hybrid AARNN model for these ten countries including world are displayed in Table 7 and 8 (A-5 & A-6) respectively.

**Table 5:**
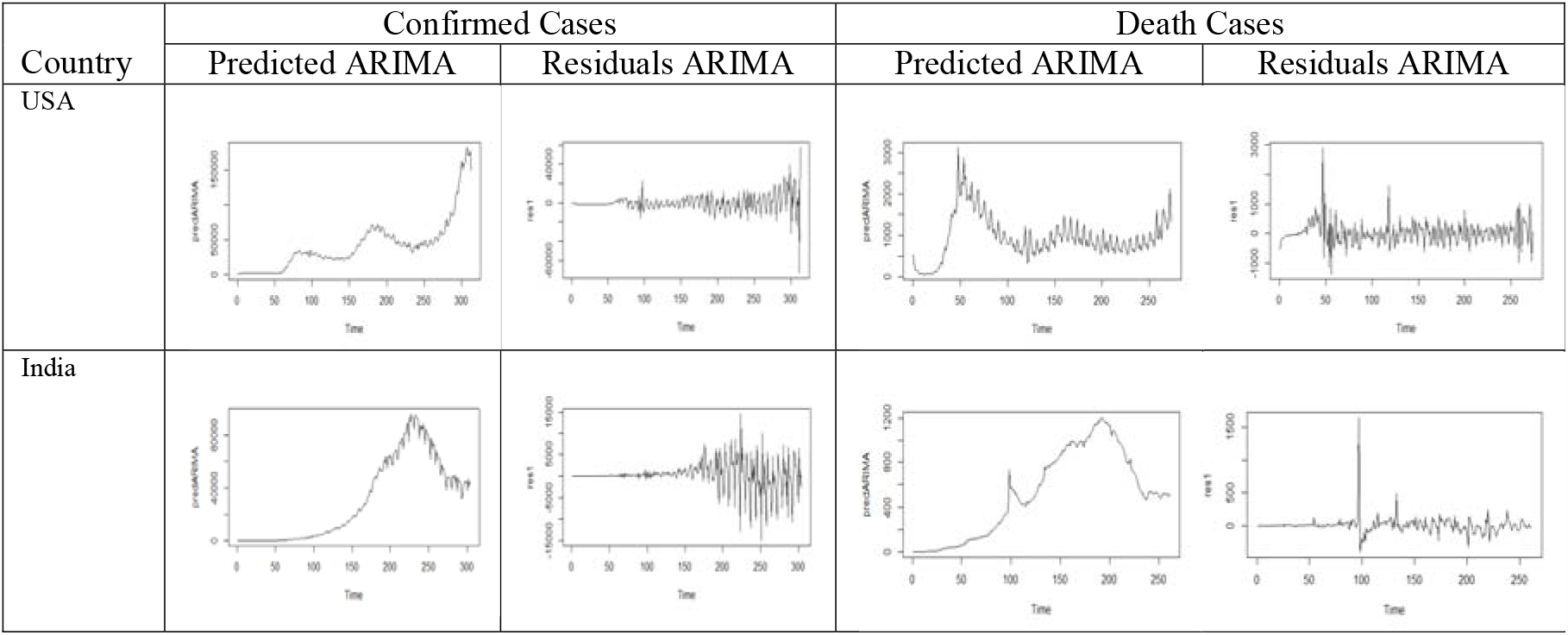
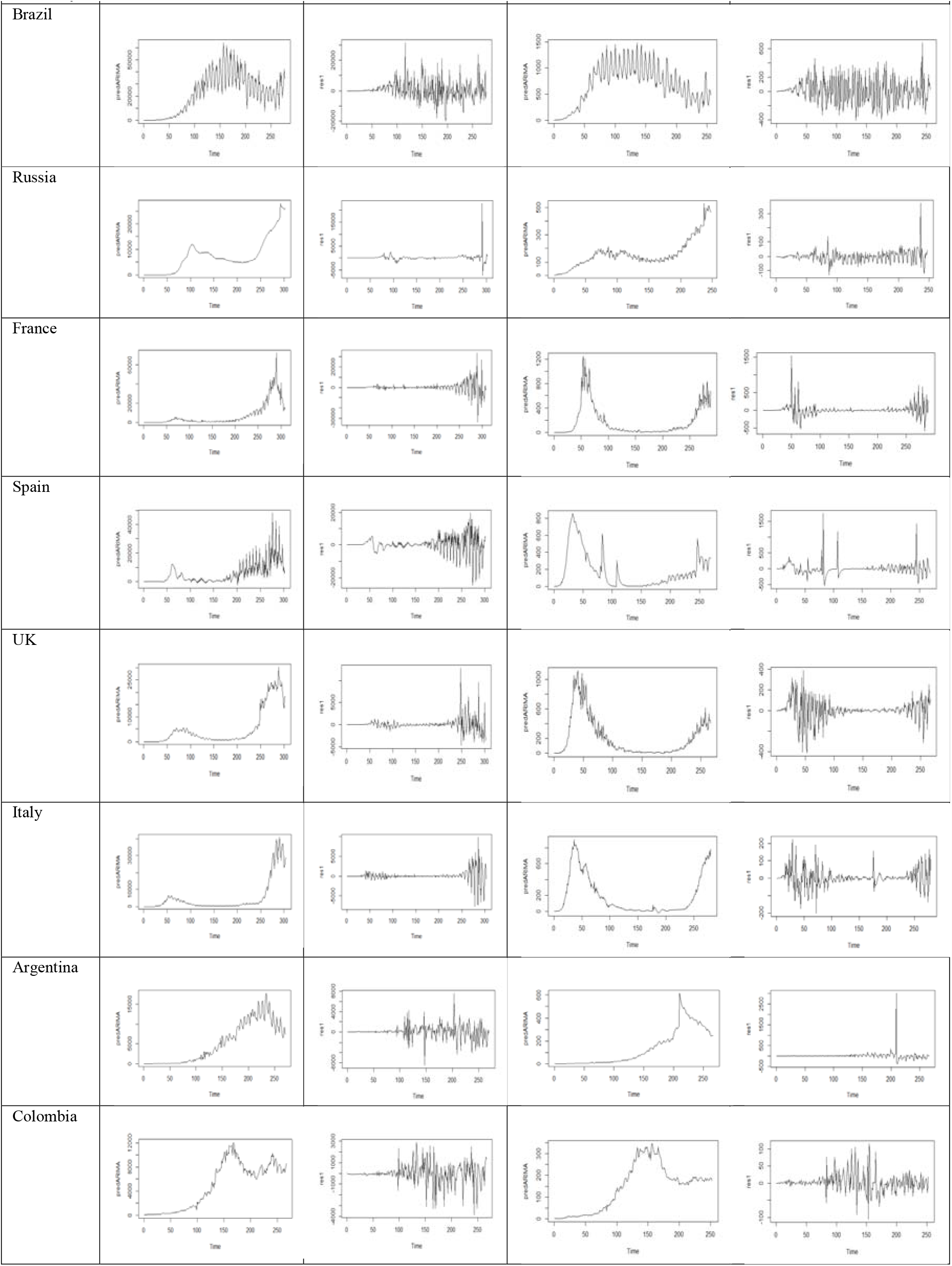
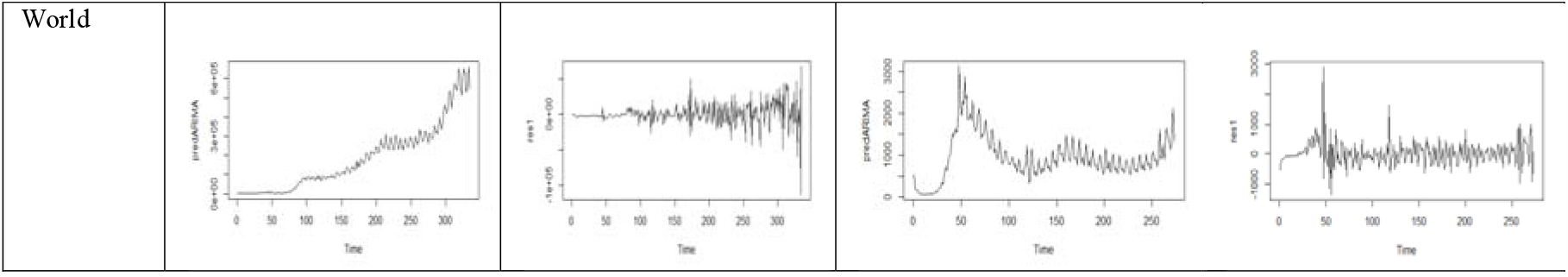
Plots of predicted and residuals of COVID-19 confirmed cases and death cases using ARIMA model for USA, India, Brazil, Russia, France, Spain, UK, Italy, Argentina, Colombia and World

**Table 6:**
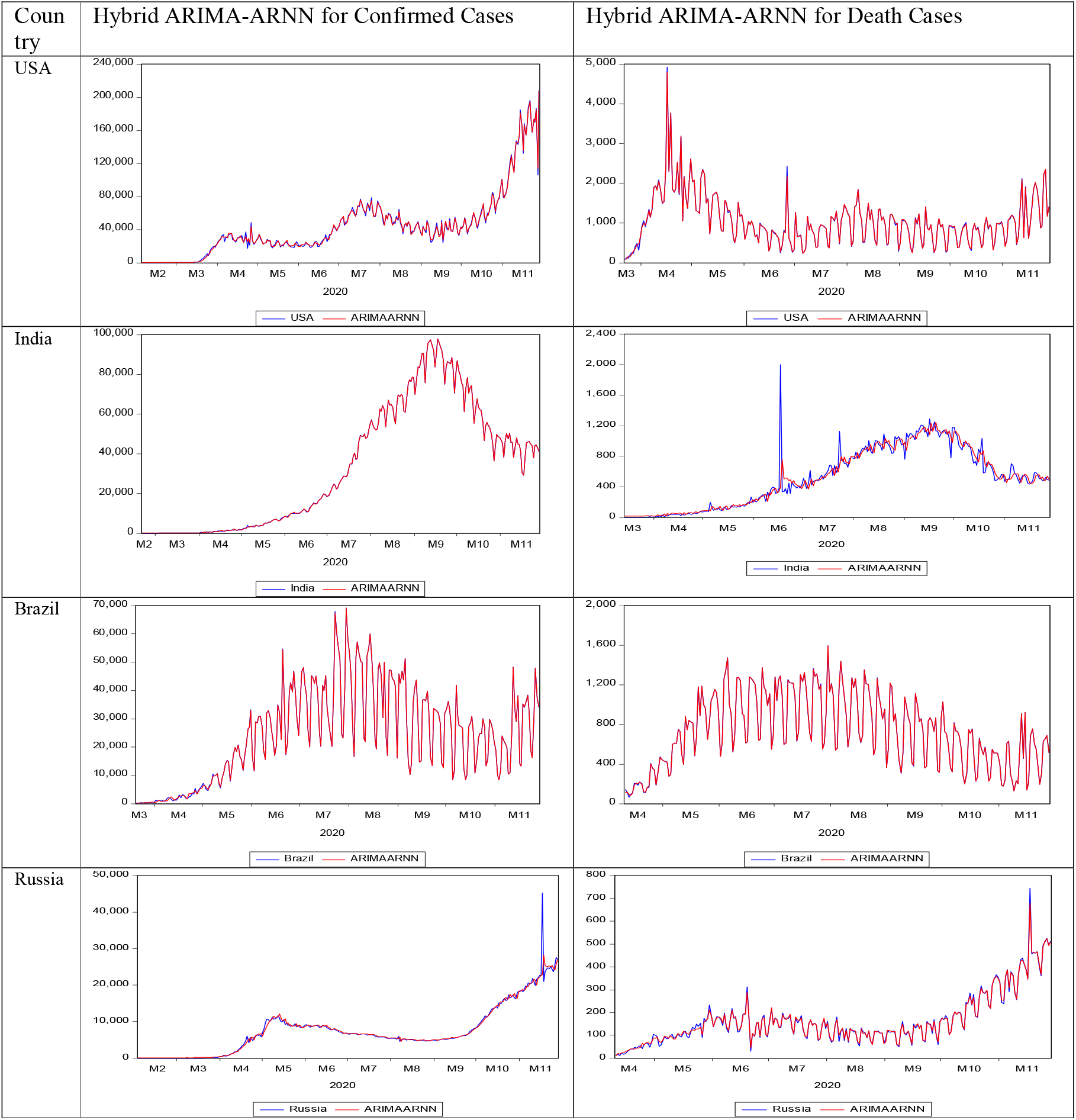
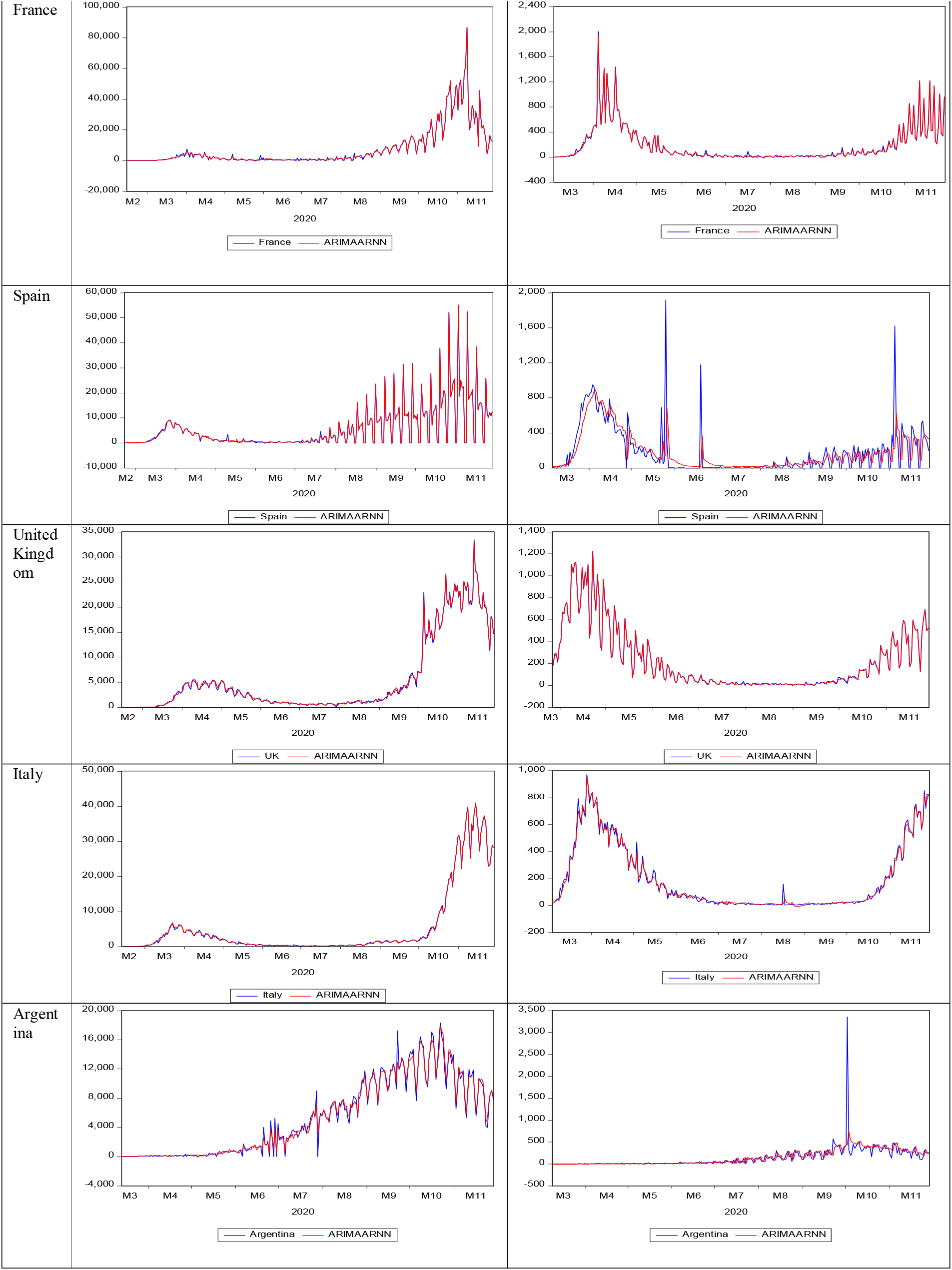
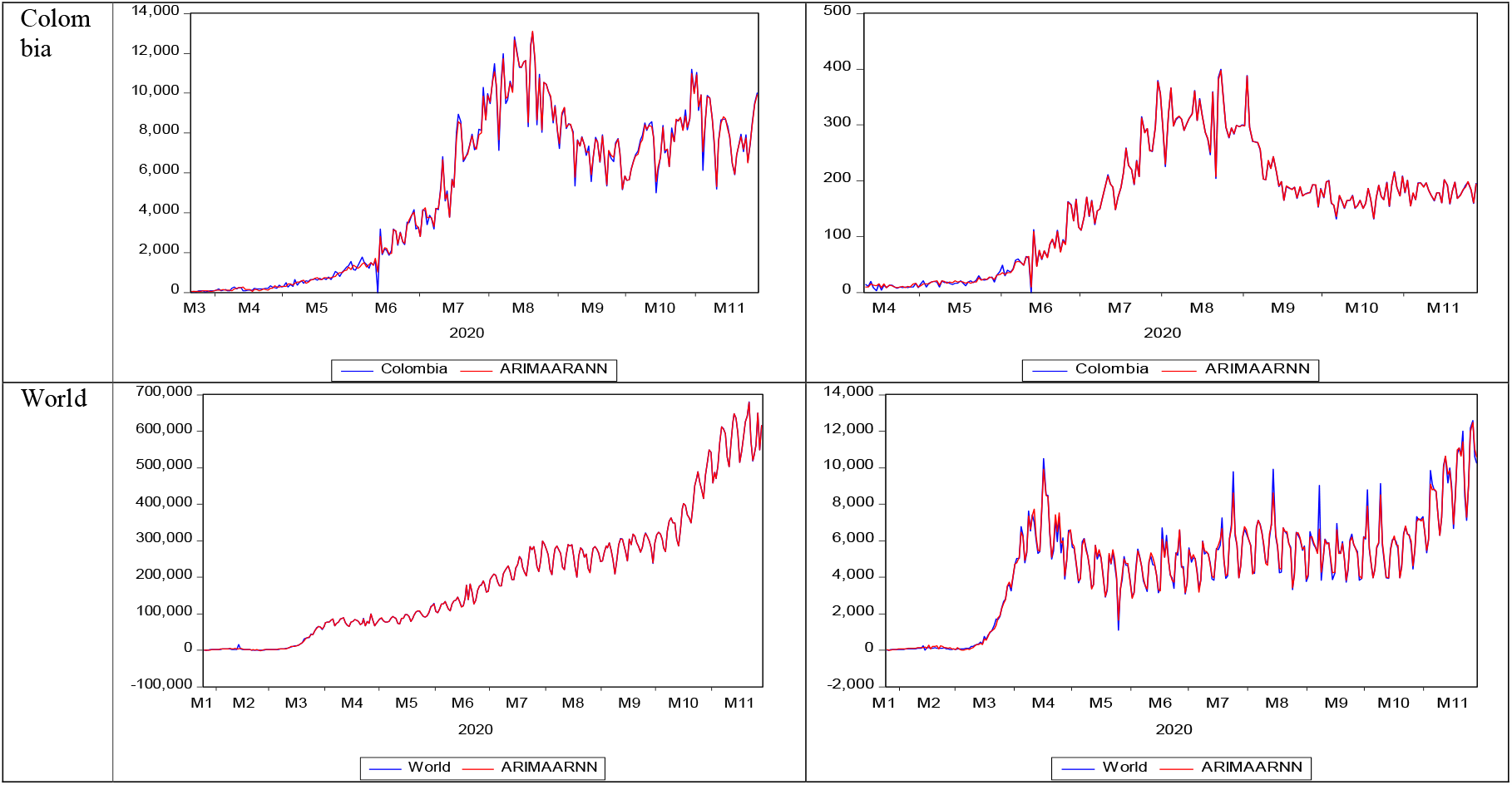
Actual and Predicted values of COVID-19 confirmed cases and death cases using hybrid ARIMA-ARNN (AARNN) model for USA, India, Brazil, Russia, France, Spain, UK, Italy, Argentina, Colombia and World

**Table 7:**
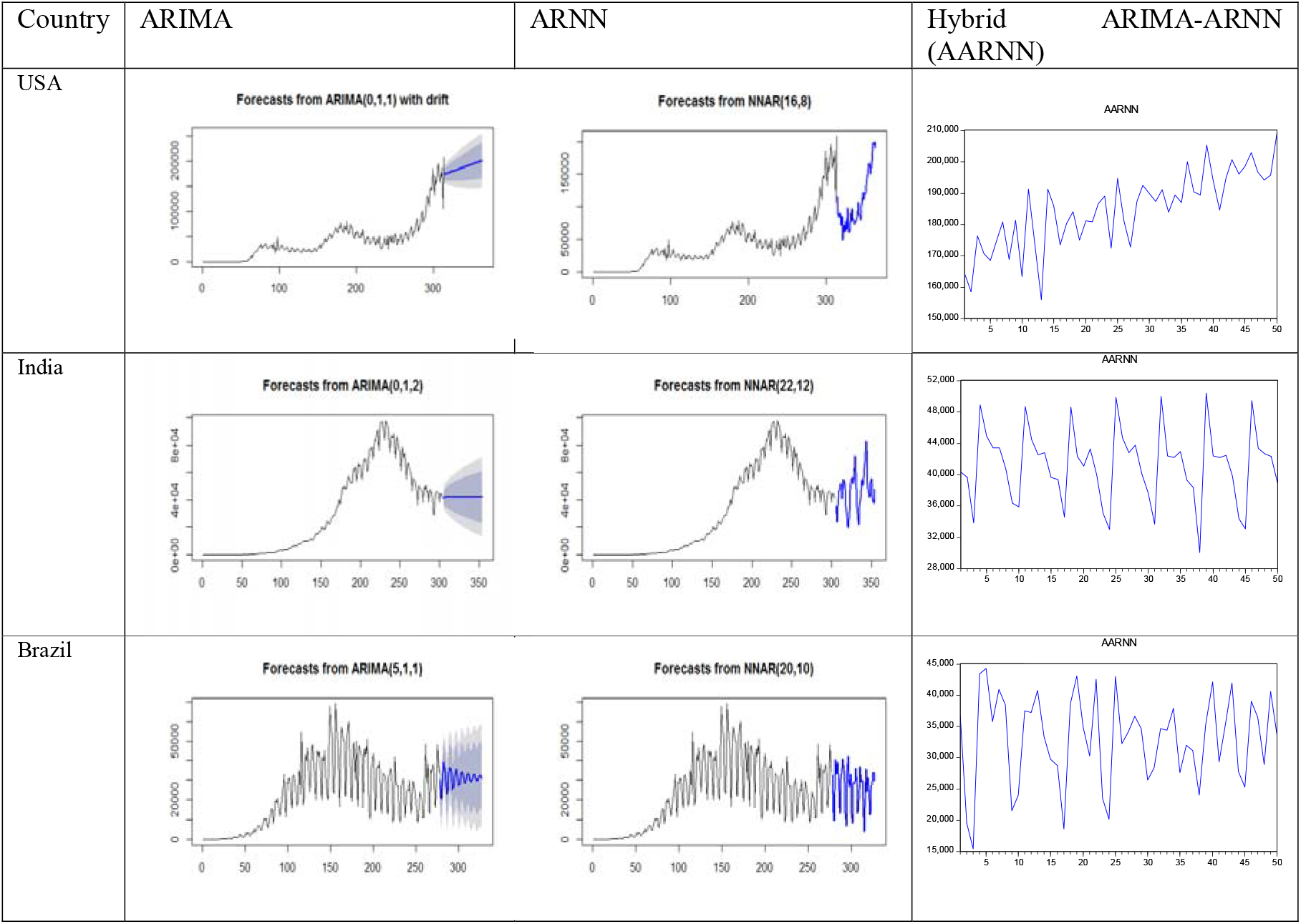
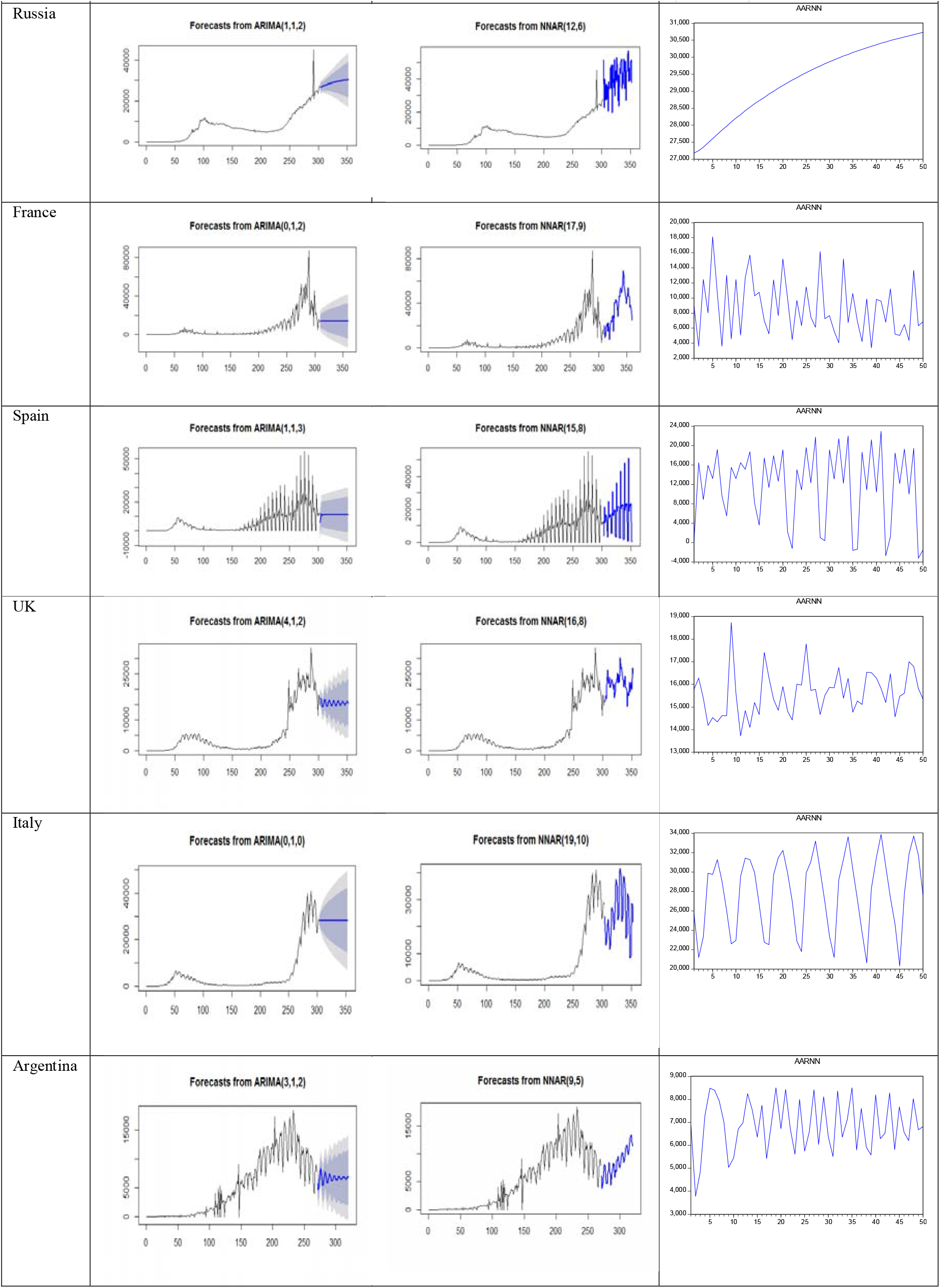
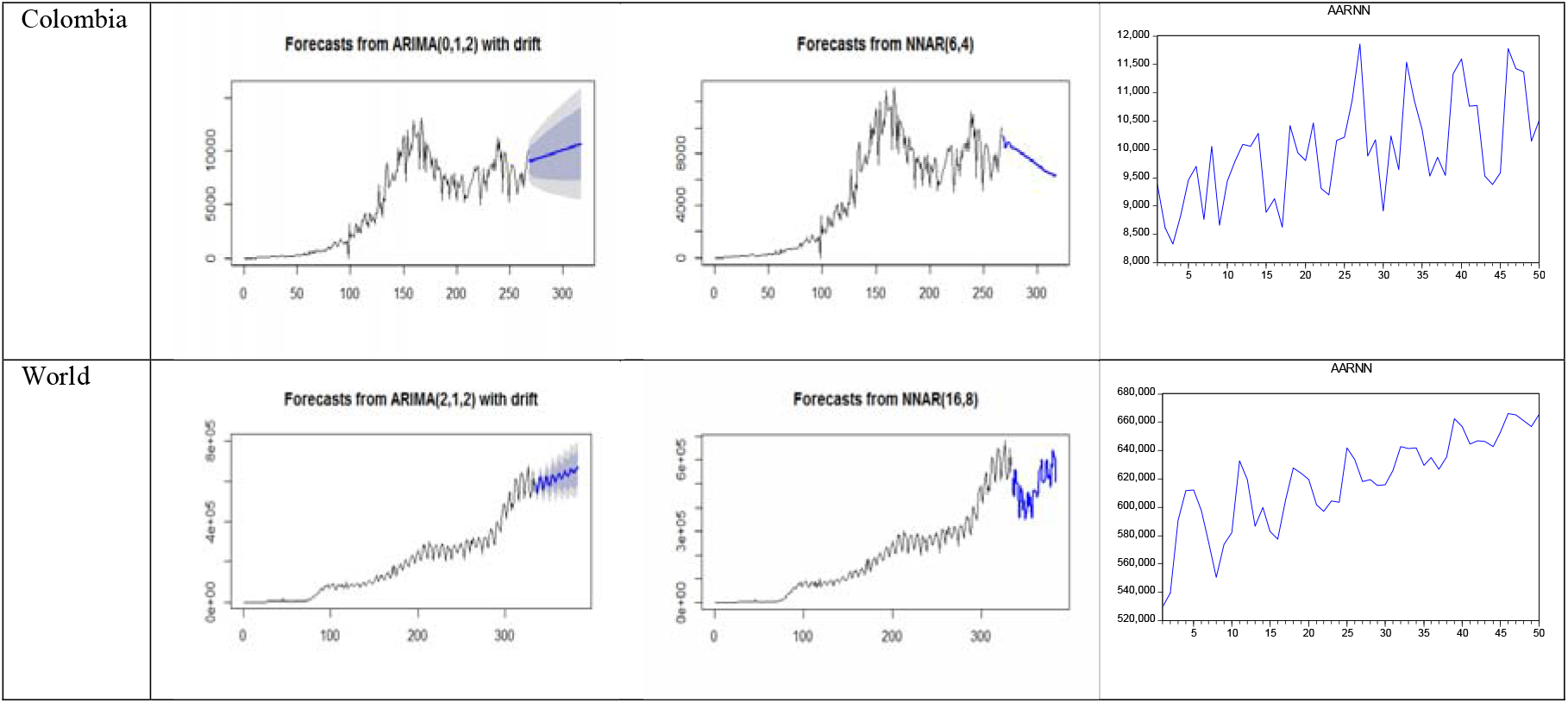
Real-time out of sample forecasts (50 days ahead) of COVID-19 confirmed cases using different forecasting models for USA, India, Brazil, Russia, France, Spain, UK, Italy, Argentina, Colombia and World

**Table 8:**
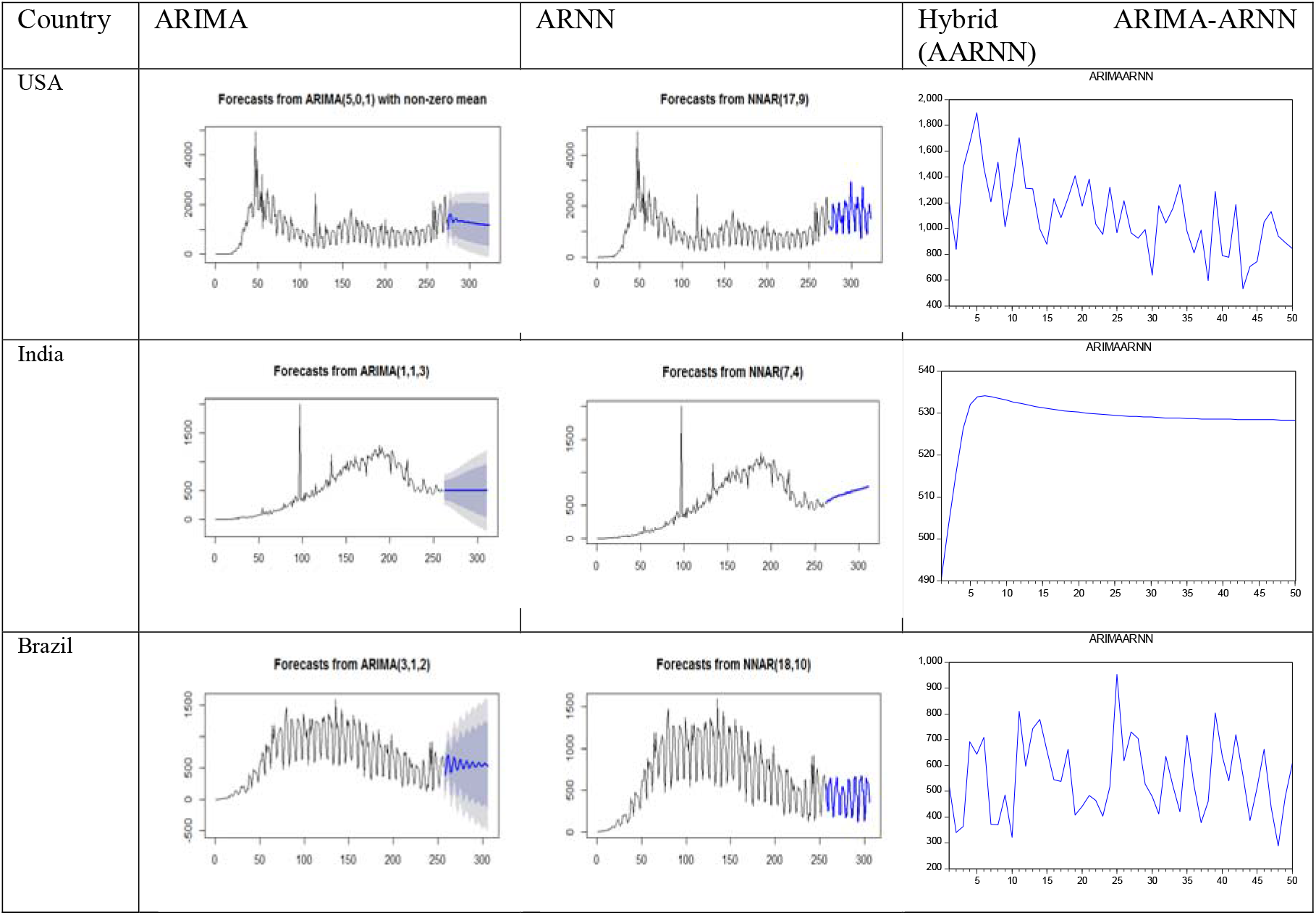
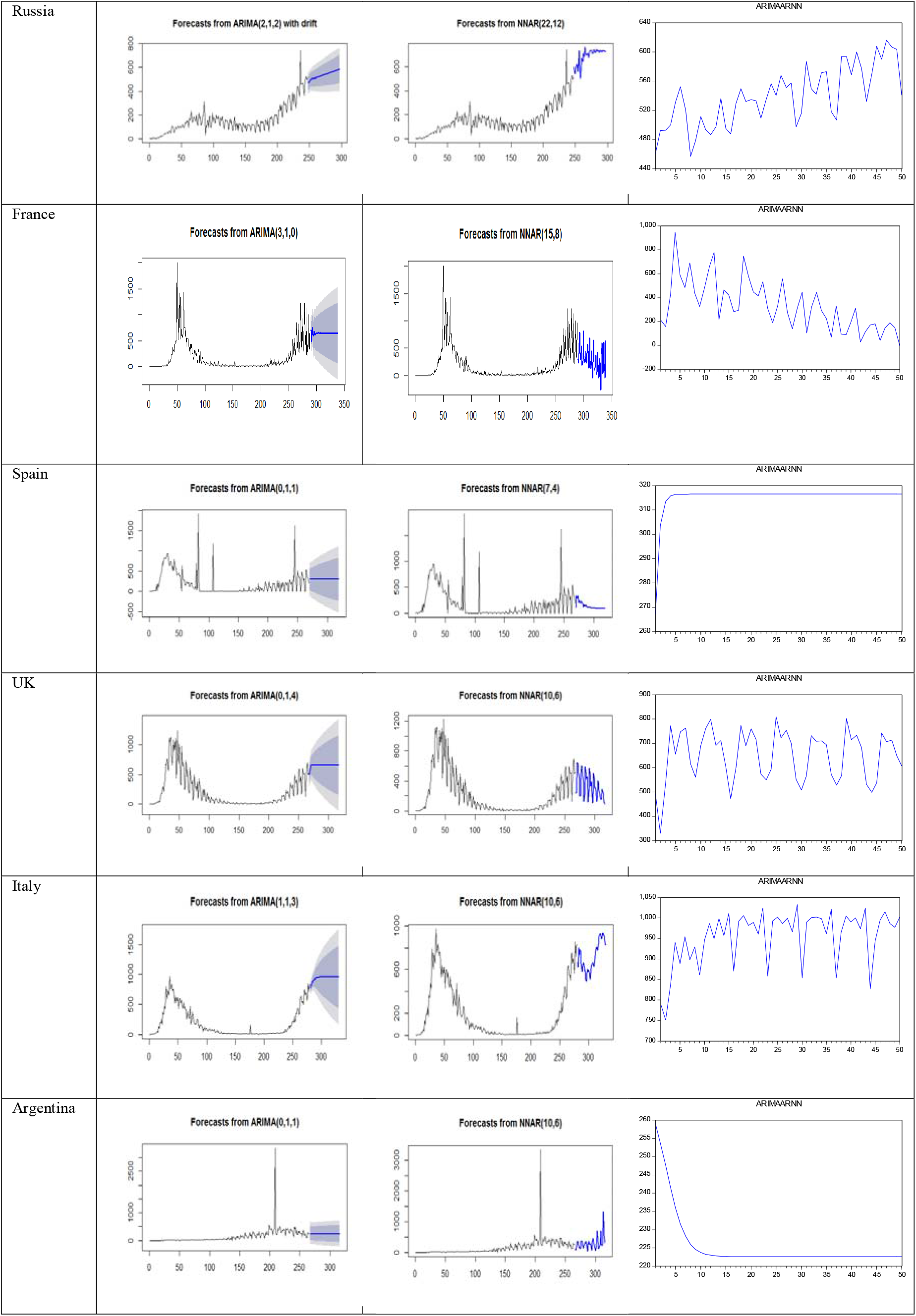
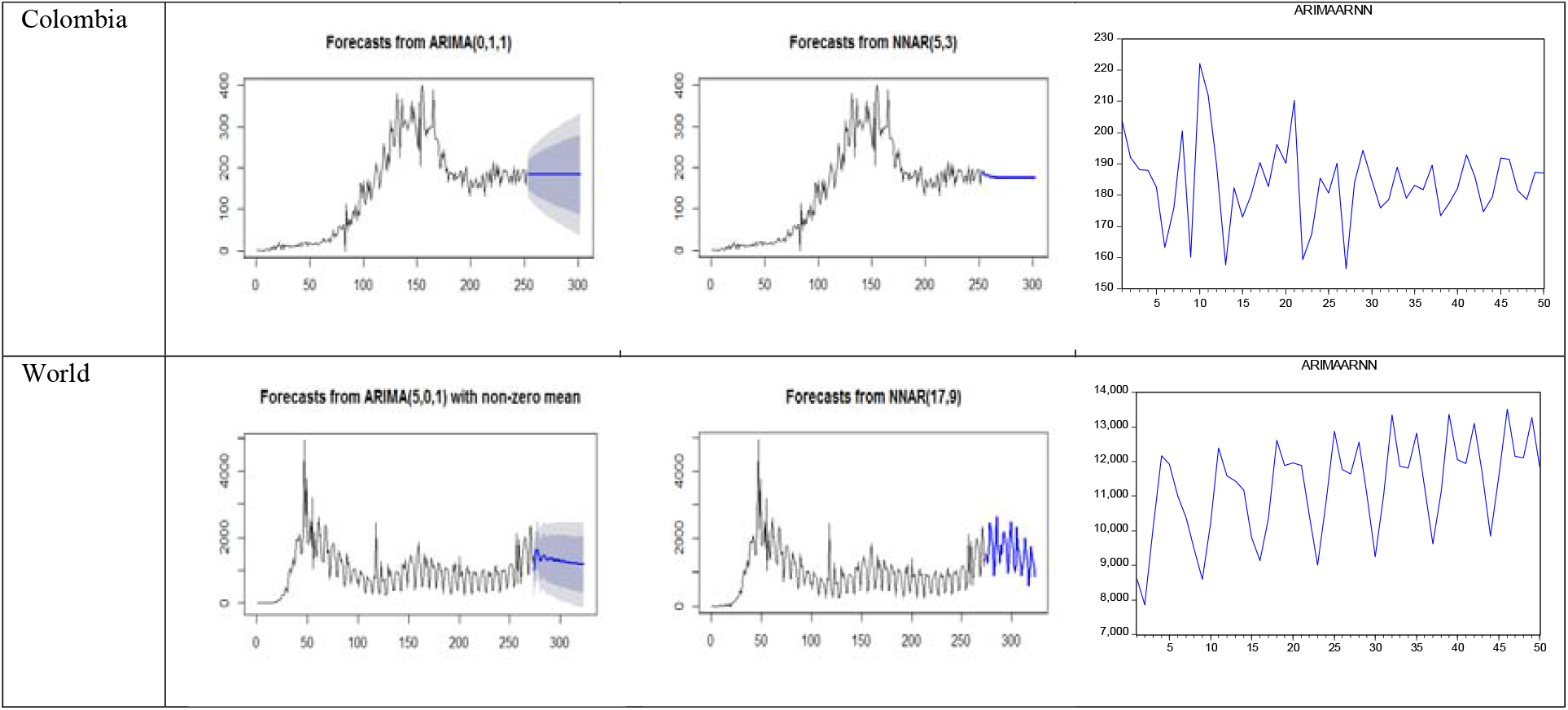
Real-time out of sample forecasts (50 days ahead) of COVID-19 death cases using different forecasting models for USA, India, Brazil, Russia, France, Spain, UK, Italy, Argentina, Colombia and World

| Country | ARIMA | ARNN | Hybrid<br>(AARNN) |
| --- | --- | --- | --- |
| USA | Forecasts from ARIMA(5,0,1) with non-zero mean | Forecasts from NNAR(17,9) | ARIMAARNN |
| India | Forecasts from ARIMA(1,1,3) | Forecasts from NNAR(7,4) | ARIMAARNN |
| Brazil | Forecasts from ARIMA(3,1,2) | Forecasts from NNAR(18,10) | ARIMAARNN |
| Russia | Forecasts from ARIMA(2,1,2) with drift | Forecasts from NNAR(22,12) | ARIMAARNN |
| France | Forecasts from ARIMA(3,1,0) | Forecasts from NNAR(15,8) | ARIMAARNN |
| Spain | Forecasts from ARIMA(0,1,1) | Forecasts from NNAR(7,4) | ARIMAARNN |
| UK | Forecasts from ARIMA(0,1,4) | Forecasts from NNAR(10,6) | ARIMAARNN |
| Italy | Forecasts from ARIMA(1,1,3) | Forecasts from NNAR(10,6) | ARIMAARNN |
| Argentina | Forecasts from ARIMA(0,1,1) | Forecasts from NNAR(10,6) | ARIMAARNN |
| Colombia | Forecasts from ARIMA(0,1,1) | Forecasts from NNAR(5,3) | ARIMAARNN |
| World | Forecasts from ARIMA(5,0,1) with non-zero mean | Forecasts from NNAR(17,9) | ARIMAARNN |

For comparison purposes, we applied traditional and advanced individual models like ARIMA, ARNN, ANN, WBF along with the hybrid models like ARIMA-ANN and WARIMA models (Chakraborty et al., 2020) the non-linearity, non-stationarity and non- Gaussian COVID-19 confirmed and death cases datasets of these ten countries including world respectively. All the experimental results are reported in Table 9. The performance of the proposed hybrid AARNN model is superior as compared to all the traditional, advanced individual and hybrid models under study. In comparison to other individual and hybrid models, nine for confirmed cases and seven for death cases out of eleven datasets of COVID- 19 respectively, our proposed hybrid AARNN model outperformed all the hybrid, traditional and advanced individual models in the significant edge. The consistency and adequacy in experimental results empirically approves the same. Thus, the efficacy of the proposed methodology of the proposed hybrid model is experimentally validated.

**Table 9:** Quantitative measures of performance for different forecasting models on eleven time series (training data sets only) of COVID-19 confirmed cases and death cases for USA, India, Brazil, Russia, France, Spain, UK, Italy, Argentina, Colombia and World

| Confirmed Cases |  |  |  |  |  |  |  |  |
| --- | --- | --- | --- | --- | --- | --- | --- | --- |
| Country | Performance | ARIMA | ARNN | ANN | WBF | ARIMA-ANN | WARIMA | AARNN |
| USA | RMSE | 9419.69 | 2104.26 | 7852.68 | 11952.24 | 9276.98 | 8496.16 | **1832.83** |
|  | MAE | 5713.55 | 1441.45 | 4786.45 | 6214.82 | 5573.66 | 5211.76 | **1337.14** |
| India | RMSE | 3356.65 | 384.99 | 3110.26 | 3544.27 | 3002.94 | 2663.07 | **149.11** |
|  | MAE | 1899.34 | 293.79 | 1816.70 | 2183.67 | 1734.80 | 1592.87 | **85.29** |
| Brazil | RMSE | 6532.36 | 608.24 | 6374.66 | 7554.62 | 6354.61 | 6667.63 | **231.97** |
|  | MAE | 4281.56 | 371.82 | 4067.49 | 5335.20 | 4267.18 | 4379.48 | **128.17** |
| Russia | RMSE | 1521.95 | **348.41** | 1380.25 | 2009.50 | 1364.49 | 1698.21 | 1431.09 |
|  | MAE | 490.39 | **209.29** | 418.88 | 793.73 | 364.01 | 471.63 | 406.77 |
| France | RMSE | 5085.97 | 692.29 | 3315.97 | 5127.78 | 5017.16 | 5722.36 | **453.53** |
|  | MAE | 2263.08 | 415.79 | 1749.46 | 2404.33 | 2339.38 | 2449.29 | **281.87** |
| Spain | RMSE | 5540.08 | 499.48 | 2843.82 | 5959.65 | 4965.75 | 5791.24 | **315.82** |
|  | MAE | 3310.28 | 335.48 | 1675.20 | 3585.93 | 3036.40 | 3242.51 | **186.86** |
| UK | RMSE | 1485.57 | **257.35** | 1365.60 | 1806.59 | 1343.56 | 1603.35 | 292.20 |
|  | MAE | 663.77 | **176.10** | 636.86 | 872.75 | 643.30 | 794.82 | 194.38 |
| Italy | RMSE | 1543.97 | 198.88 | 1134.29 | 1808.85 | 760.11 | 1764.39 | **191.64** |
|  | MAE | 669.20 | 133.61 | 546.17 | 778.34 | 391.19 | 740.37 | **133.71** |
| Argentina | RMSE | 1348.16 | 730.81 | 1156.72 | 1551.44 | 1332.41 | 1408.51 | **586.65** |
|  | MAE | 796.25 | 422.52 | 686.77 | 989.76 | 791.59 | 840.03 | **357.81** |
| Colombia | RMSE | 940.37 | 761.52 | 863.32 | 1076.17 | 935.04 | 1062.37 | **164.40** |
|  | MAE | 584.57 | 472.22 | 559.24 | 691.78 | 592.03 | 681.38 | **106.60** |
| World | RMSE | 15882.62 | 5960.31 | 20129.09 | 29358.84 | 13000.84 | 15849.38 | **864.36** |
|  | MAE | 10107.56 | 4079.81 | 13411.11 | 15705.79 | 8508.95 | 10445.66 | **320.72** |
| Death Cases |  |  |  |  |  |  |  |  |
| Country | Performance | ARIMA | ARNN | ANN | WBF | ARIMA-ANN | WARIMA | AARNN |
| USA | RMSE | 407.57 | 68.77 | 332.16 | 377.55 | 326.85 | 413.23 | **27.82** |
|  | MAE | 280.05 | 41.53 | 214.06 | 254.89 | 228.69 | 295.85 | **15.67** |
| India | RMSE | 132.63 | **89.97** | 125.81 | 139.63 | 126.23 | 147.95 | 127.44 |
|  | MAE | 59.22 | **47.91** | 54.56 | 68.45 | 53.78 | 67.36 | 56.58 |
| Brazil | RMSE | 182.04 | 17.35 | 151.28 | 189.57 | 135.63 | 151.30 | **4.46** |
|  | MAE | 140.64 | 11.40 | 106.25 | 142.92 | 103.33 | 108.10 | **2.34** |
| Russia | RMSE | 43.13 | **4.19** | 38.82 | 51.29 | 41.02 | 43.95 | 8.35 |
|  | MAE | 28.52 | **1.99** | 25.54 | 33.09 | 27.47 | 29.94 | 5.52 |
| France | RMSE | 166.09 | 17.54 | 125.57 | 167.66 | 140.30 | 196.74 | **13.84** |
|  | MAE | 73.38 | 11.68 | 53.77 | 79.23 | 62.61 | 94.58 | **8.50** |
| Spain | RMSE | 202.58 | **130.04** | 136.47 | 217.53 | 201.60 | 232.43 | 197.72 |
|  | MAE | 98.84 | **62.11** | 70.97 | 115.94 | 100.43 | 111.27 | 94.37 |
| UK | RMSE | 106.68 | 25.76 | 84.23 | 116.01 | 104.94 | 113.39 | **6.21** |
|  | MAE | 64.57 | 16.99 | 50.96 | 74.50 | 62.93 | 67.41 | **4.05** |
| Italy | RMSE | 52.79 | 21.30 | 56.91 | 73.32 | 52.78 | 56.91 | **19.96** |
|  | MAE | 31.03 | 13.83 | 31.79 | 40.16 | 31.40 | 34.55 | **12.90** |
| Argentina | RMSE | 200.46 | **40.96** | 192.44 | 216.70 | 193.26 | 218.56 | 194.96 |
|  | MAE | 54.64 | **25.36** | 53.64 | 67.84 | 54.17 | 61.71 | 57.45 |
| Colombia | RMSE | 27.18 | 24.92 | 24.23 | 31.62 | 26.60 | 28.55 | **23.58** |
|  | MAE | 17.11 | 14.83 | 14.20 | 19.75 | 16.98 | 18.25 | **1.29** |
| World | RMSE | 892.20 | 455.11 | 727.49 | 1055.20 | 786.25 | 910.57 | **245.82** |
|  | MAE | 609.92 | 261.79 | 471.77 | 722.90 | 512.99 | 605.25 | **134.04** |

## 5. Practical implications and discussion

In this study, a novel hybrid AARNN model is established that performs considerably well for both confirmed and death cases of COVID-19 forecasting for the top ten countries namely USA, India, Brazil, Russia, France, Spain, UK, Italy, Argentina, Colombia including world respectively. The proposed novel hybrid AARNN model filters linearity using the ARIMA model and on the whole, it can explain the linearity, nonlinearity and nonstationary tendencies present in both the confirmed cases and death cases of COVID-19 data sets of ten different countries including world better as compared to the traditional, advanced individual and hybrid models. The proposed hybrid AARNN model also yields better forecast accuracy than various traditional, advanced individual and hybrid models like ARIMA, ARNN, ANN, WBF, ARIMA-ANN and WARIMA for nine and seven out of eleven data sets of both confirmed and death cases of COVID-19 respectively considered in this study. The proposal will be useful in decision and policy makings for government officials and policymakers to allocate adequate health care resources for the coming days in responding to the crisis. Although any epidemics time series can oscillate heavily because of various epidemiological factors and these fluctuations are generally challenging to be fittingly captured for precise forecasting. However, the proposed novel hybrid model may still predict with better accuracy. LTM of fifty days ahead out of sample forecasts of both confirmed and death cases of COVID-19 separately are provided for all the ten countries including world. The proposed model can be used as a premature word of warning to struggle against the COVID-19 global pandemic. A list of suggestions based on the results of the real-time forecasts using a novel data-driven hybrid methodology is presented below:

1. Since we presented a real-time forecast system unlike ex-post analysis, thus one can regularly update both the actual daily confirmed and death cases and update the predictions, just like it happens in weather forecasting or other epidemics forecasting.
2. The forecasts on the whole follow oscillating behaviour for the out of sample forecast of 50 days ahead respectively for the ten different countries namely USA, India, Brazil, Russia, France, Spain, UK, Italy, Argentina, Colombia including world and reflect the impact of the broad spectrum of social distancing, wearing masks, containment zone, lock down, shutdown, quarantine and sanitizing properly measures implemented by the governments, which likely promoted stable the epidemic.
3. The LTM out of sample forecasts of confirmed cases show oscillatory behaviour with upward trend and don’t show any stiff decay sooner except France and Spain and stiff amplify behaviour with upward trend for Russia. The LTM out of sample forecasts of death cases show oscillatory behaviour with both upward and downward trend and don’t show any stiff decay sooner except Russia. India, Spain and Argentina don’t show any oscillatory behavior. All the six countries including world except France, Spain, UK and Italy are going to face unlike uplifts in the number of new confirmed cases of COVID-19 pandemic.
4. Followed by the LTM out of sample forecasts reported in this paper, the wearing masks, sanitization, immunity recreation, social distancing, containment zone, self precaution etc. can be implemented accordingly to tackle the uncertain and robust situations of COVID- 19 global pandemic.

## 6. Conclusions

From the data-driven experimental analysis, it is established that all the traditional and advanced individual and hybrid models like ARIMA, ARNN, ANN, WBF, ARIMA-ANN and WARIMA models are unable to completely capture the behaviour of the nonlinearity, non-stationarity and non-Gaussian of the stochastic time series datasets containing intrinsic random shock or moving average component. This indicates the need to develop a novel hybrid model with AARNN to predict the subsequent both the confirmed and death cases of COVID-19 outbreaks accurately and respond to global pandemics more powerfully.

## Data Availability

https://ourworldindia.org/coronavirus

## Abbreviations

COVID-19: Coronavirus Disease 2019
ARIMA: Auto Regressive Integrated Moving Average
ARNN: Auto Regressive Neural Network
AARNN: ARIMA-ARNN
LTM: Long Term Memory
ANN: Artificial Neural Network
WBF: Wavelet Based Forecasting
USA: United States of America
UK: United Kingdom
JB: Jarque Bera test
RMSE: Root Mean Square Error
MAE: Mean Absolute Error
ACF: Auto- correlation function
PACF: Partial Auto-correlation function
AIC: Akaike Information Criterion
BIC: Bayesian Information Criterion
WARIMA: WBF-ARIMA
AR: Auto Regressive
MA: Moving Average

## Appendix

### A-1

### A-2

### A-3

### A-4

### A-5

### A-6

## Footnotes

1 https://ourworldindia.org/coronavirus

